# Predictors of statin intolerance among statin users in primary care using real-world data

**DOI:** 10.64898/2026.02.23.26346866

**Authors:** Shagoofa Rakhshanda, Siaw-Teng Liaw, Joel Rhee, Kerry-Anne Rye, Jitendra Jonnagaddala

## Abstract

**Objective:** The objective of this study was to identify the demographic and clinical predictors of statin intolerance among patients prescribed statins for the primary prevention of cardiovascular disease during the first two years following the first recorded prescription, using real-world data.

**Methods:** This study used the Electronic Practice Based Research Network Linked Dataset. An operational definition based on muscle symptoms and creatine kinase levels was applied to identify patients with statin intolerance. Descriptive and multivariate logistic regression analyses were performed using R software. All variables identified a priori as potential predictors were included in the initial model, and backward stepwise selection using the Akaike Information Criterion was performed across the imputed datasets to identify a parsimonious final model.

**Results:** Overall, 4,016 patients accounting for 60,873 visits were included in the analysis. About 3.5% of patients were statin intolerant. After adjusting for all other variables, higher odds of statin intolerance were observed among female or other gender patients (AOR 1.5, 95% CI 1.0 – 2.2), patients residing in Socio-Economic Indexes for Australia (SEIFA) area based socioeconomic advantage and disadvantage (IRSAD) category 8 or 9 (AOR 3.4, 95% CI 2.0 – 5.8), and patients with three or more selected comorbidities (AOR 6.4, 95% CI 2.2 – 18.3). A similar direction of associations was seen in the final model.

**Conclusion:** Gender, IRSAD, and the number of selected comorbidities were associated with statin intolerance. However, these findings should be interpreted in the context of the operational EHR based definition of statin intolerance. Further validation in other populations and settings is required before these predictors can be considered for clinical application.

## 1. Introduction

Statins are a group of lipid-modifying drugs that are prescribed as primary and secondary prevention of cardiovascular diseases (CVDs) in Australia (1). They are commonly used as the first line of pharmacological therapy for CVD risk management, and are prescribed to 30% of Australian patients aged more than 55 years (2, 3). Although statins are normally quite well tolerated, statin intolerance can occur partially in about 15% patients, and complete statin intolerance can occur in about 5% patients (4, 5). Statin intolerance is defined differently by different scientific groups, trialists, and societies (6). However, it is commonly known as the inability to tolerate statin therapy due to adverse effects, most notably statin-associated muscle symptoms (SAMS) (6, 7). Symptoms of statin intolerance include weakness, muscle aches (myalgia), or cramps, myopathy, memory loss and rhabdomyolysis. Some of the other symptoms include sleep disturbances, peripheral neuropathy, diabetes mellitus, erectile dysfunction, and increased creatinine kinase (4, 5, 8). Statin intolerance may effect statin adherence which can result in increased risk of CV events (9, 10).

The literature suggests a range of demographic and clinical factors that affect statin intolerance, such as age, gender, statin intensity, and comorbidities (11, 12). These factors are the key targets for any intervention used to manage statin intolerance. It is found that older age, female gender, prescription of higher intensity statins and burden of more than one comorbidity are strongly associated with statin intolerance (4, 13). However, these findings may vary based by study population and methodology. Studies have also found that pharmacogenetic and immunogenetic factors, such as LILRB5 (Leukocyte immunoglobulin-like receptor subfamily B member 5) variants and specific HLA (Human Leukocyte Antigen) alleles, are also potential predictors for statin intolerance (14, 15, 16). Additional reviews discuss clinical risk factors such as hypothyroidism, vitamin D deficiency, renal impairment, and interactions with CYP3A4 (Cytochrome P450, family 3, subfamily A, polypeptide 4) inhibitors, all of which can contribute to muscle-related adverse events or mimic statin intolerance (12, 17). Diabetes mellitus is also a factor that may lead to statin intolerance (18).

A substantial contributor to reported statin intolerance is the nocebo/drucebo effect, whereby patient expectations and awareness may influence the reporting of muscle or other symptoms attributed to statin therapy (6, 19, 20, 21). Negative information about statins from the media, close associates, or other sources may contribute to these expectations (22, 23). However, patient expectations, beliefs, and symptom attribution are not routinely captured in structured electronic health records (EHR) data from primary care practices. They are usually documented as text information in the EHR and not extracted for privacy reasons. Therefore, direct assessment of this effect was outside the scope of this study. The present study focused on demographic and clinical factors that could be identified from routinely collected EHR data. The potential contribution of unmeasured patient perceptions should be considered when interpreting predictors of statin intolerance.

Statin intolerance is a clinically important barrier to effective lipid management. Identifying predictors of statin intolerance may help identifying patients who could benefit from closer monitoring following statin initiation. The potential value of a prediction model is therefore not to replace the clinical assessment of SAMS or to determine treatment changes before symptoms occur. Rather, information available in the EHR could potentially be used to identify patients at higher risk and support earlier follow-up, enquiry about symptoms, review of potentially modifiable risk factors, and discussion about statin therapy. If symptoms subsequently develop, established management strategies such as assessment of alternative causes, dose modification, statin rechallenge, or consideration of alternative therapies can then be applied according to the individual clinical context (24, 25, 26). However, any prediction model would require adequate predictive performance and external validation before being considered for clinical use.

Therefore, the objective of this study was to identify the demographic and clinical predictors of statin intolerance among patients prescribed statins for the primary prevention of CVD during the first two years following their first recorded statin prescription, using routinely collected general practice EHR data. The study aimed to assess whether factors available within the EHR could contribute to the identification of patients at increased risk of statin intolerance and potentially inform more targeted monitoring and intervention in primary care.

## 2. Methods

### 2.1 Data source

The dataset used for this study was the Electronic Practice Based Research Network (ePBRN) Linked Dataset, which is derived from general practice EHRs in southwest Sydney (SWS) local health district (LHD). The dataset contains information about 249,345 patients spread across 11 general practitioner sites, who had at least one visit between 2012 and 2019 (27, 28).

There are 76 tables in the ePBRN dataset, of which 39 tables were sourced from Best Practice, and 37 were sourced from Medical Director. This study required a selection of data on statin intolerance from these tables to identify patients with relevance and precision. For this, 6 tables (Patients, Visits, Visit Reason, Current Rx, Careplan Goal and Past History) and 5 tables (Patients, History, Diagnosis, Consultations and Prescription) were selected to extract data on patient diagnosis, conditions, visit details, and demographics from Best Practice and Medical Directors respectively.

### 2.2 Selection criteria

The ePBRN dataset contained general practice EHR records collected between 2012 and 2019, which defined the available study period. Patients with a recorded statin prescription, under any brand name, were included in this study. The statin types found in the ePBRN dataset are atorvastatin, rosuvastatin, simvastatin, pravastatin, or fluvastatin. The list of statin brand names included in the ePBRN dataset is provided in **Supplement 1**.

For this study, a Master table was developed consisting of patients with a recorded statin prescription. The index date was defined as the date of the first recorded statin prescription for each patient in the ePBRN dataset. All available EHR records before the index date were used as the lookback period to identify previously recorded statin use. No fixed run-in period was applied as the length of available EHR records varied between patients. Patients with any recorded statin prescription before the index date were excluded. Therefore, the index prescription represents the first statin prescription recorded in the available ePBRN data and may not represent the patient’s first ever use of a statin.

Patients were included if they were aged 18 years or older at the index date and had at least two years of observation following the index date. To ensure sufficient longitudinal EHR information, patients were required to meet the RACGP definition of an active patient, with at least three general practice visits for any reason during the two years observation period (29). Patients were excluded if they had no recorded gender information or birth year, were aged over 80 years at the index date, died during the observation period, or experienced a CVD event before or during the observation period.

Patients aged over 80 years were excluded because of the greater clinical complexity and variation in treatment patterns associated with advanced age (30, 31), while patients who died during the observation period did not have the complete two years of observation required for the analysis. Adults aged 18 years or older were included because statin therapy may be considered in younger adults at high cardiovascular risk (32). Patients who experienced a CVD event were excluded as a CVD event changes the indication for statin therapy and may subsequently influence treatment and prescribing patterns.

### 2.3 Outcome

Following the selection of patients who met the eligibility criteria, an operational definition was applied to identify patients with statin intolerance. As shown in **Figure 1**, patients with elevated CK, with or without recorded muscle symptoms, were classified as statin intolerant, while those without elevated CK were not classified as statin intolerant. Although statin associated muscle symptoms can occur without CK elevation, this more restrictive definition was used because the routinely collected EHR data did not consistently contain sufficient information on symptom characteristics, their temporal relationship with statin treatment, or dechallenge and rechallenge to reliably attribute muscle symptoms with normal CK to statin therapy. This approach was used to increase the specificity of case identification and reduce the potential classification of non-specific muscle symptoms such as statin intolerance.

**Figure 1:**
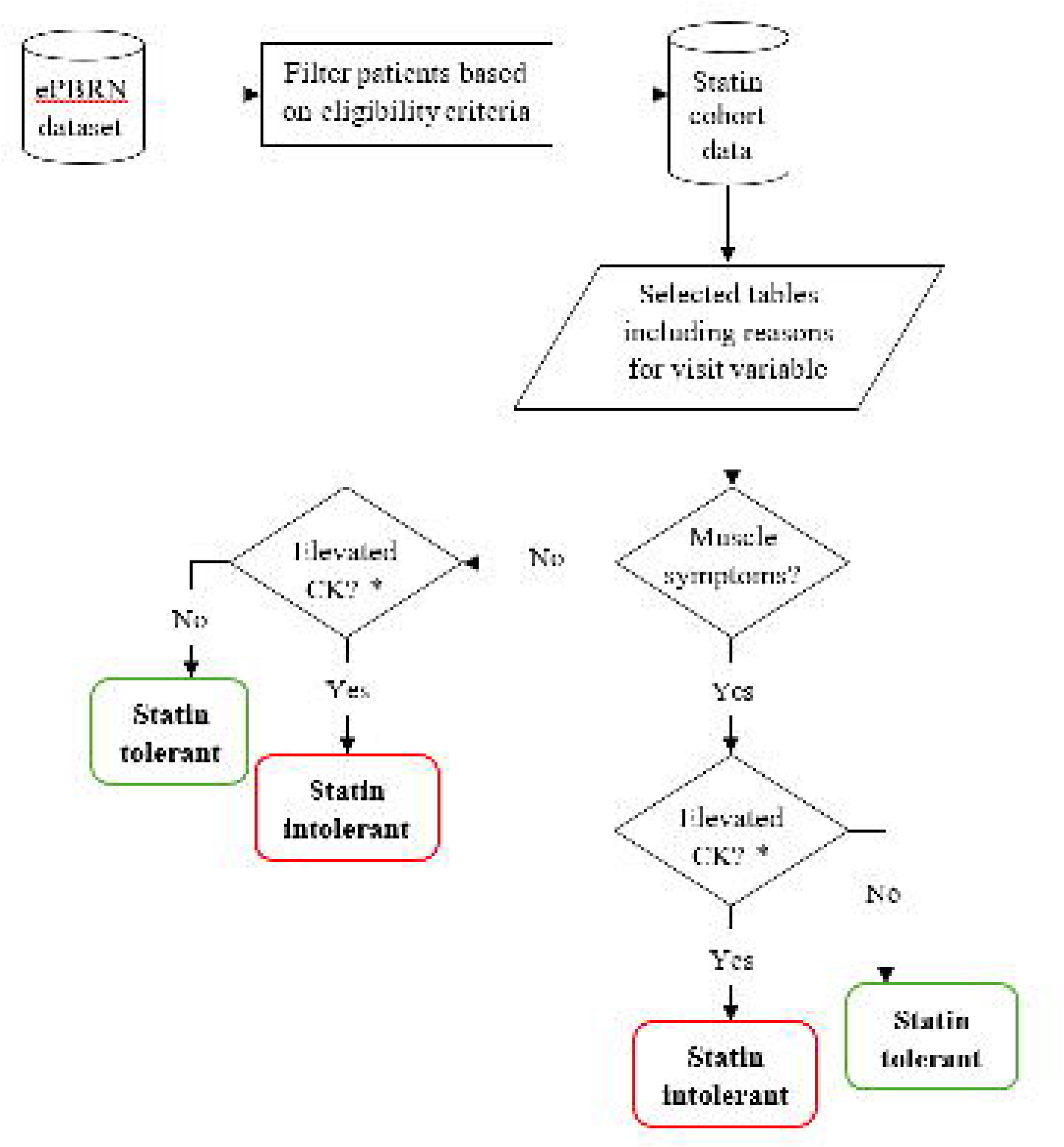
Overview of the identification of statin intolerant patients. **Notes:** ePBRN: Electronic Practice Based Research Network; CK: Creatine kinase * The threshold for creatine kinase (CK) was four times the upper limit of normal (ULN) for CK which was defined as 145–150 U/L for female patients and about 170–250 U/L for male patients

### 2.4 Covariates

The covariates for this study included patient demographics such as age (characterized as “<30”, “30-39”, “40-49”, “50-59”, “60-69” and “≥70”), gender, ethnicity, and employment status. The area-based social advantage and disadvantage index, as classified by the Socio-Economic Indexes for Australia (SEIFA) 2021 Index of Relative Socio-Economic Advantage and Disadvantage (IRSAD), based on suburbs and localities was also included as a covariate (33). Other covariates included health- and statin-related factors, such as smoking status, intensity of statin at the index date, type of statin at the index date, number of different types of statins used during the first two years, change in statin intensity between the index date and the last visit within two years, number of prescribed medications, and number of selected comorbidities. Statin intensity was classified according to **Supplement 2**.

The number of prescribed medications represented the total number of medications prescribed, including the statin. For example, a value of one represented statin use only, while a value of two represented a statin and one additional medication. Non-statin medications were grouped as anti-diabetic, anti-hypertensive, anti-coagulant, and other medications. The number of selected comorbidities represented the number of recorded conditions from the predefined categories of diabetes, hypertension, chronic respiratory disease, cancer, stroke, blood clots and Parkinson’s disease. A value of zero indicated that none of these predefined comorbidities were recorded, while a value of one indicated one recorded condition from these categories.

### 2.5 Statistical analyses

At first, descriptive analyses of statin tolerance and intolerance were performed based on all covariates and predictor variables. To identify the predictors of statin intolerance, a multivariate logistic regression analysis was performed using a backward stepwise selection approach (34, 35). All variables identified a priori as potential predictors were included in the initial model, irrespective of their univariate statistical significance or the extent of missing information. Missing information was addressed using multiple imputation by chained equations (MICE) (36, 37). The “not recorded” categories were treated as missing values, and 50 imputed datasets were generated using the available predictor variables and the outcome of statin intolerance. Multiple imputation was performed under the assumption that the data were missing at random. Regression models were fitted to the imputed datasets, and the estimates were combined using Rubin’s rules to obtain pooled odds ratios (ORs), 95% confidence intervals (CIs), and p-values. The Akaike Information Criterion (AIC) was used to guide the removal of variables. Backward stepwise selection was performed separately in each imputed dataset, and the most frequently selected model was used as the reduced final model. This model was subsequently fitted across all 50 imputed datasets, and the regression estimates were pooled using Rubin’s rules. At each step, competing models were compared and the model with the lowest AIC was preferred, resulting in a more parsimonious final model (38, 39, 40). The stability of predictor selection was further assessed using 100 bootstrap resamples of the original dataset with multiple imputation. The assumptions of logistic regression were assessed, and the associations between the selected predictors and statin intolerance were presented as adjusted ORs (AORs) and 95% CIs. All analyses were performed using R software version 4.3.2.

### 2.6 Ethical approval

This study and the use of data from ePBRN was approved by the UNSW Human Research Ethics Advisory Panel (HC230066; 23 June 2023).

## 3. Results

### 3.1 Patient selection

In the ePBRN dataset, from a total cohort of 249,345 patients accounting for 3,103,072 visits, 18,377 patients accounting for 110,147 visits were prescribed statins. After excluding patients based on the exclusion criteria (14,361 patients with 49,274 visits), 4,016 patients with 60,873 visits were included for analysis. The details of patient selection process are shown in **Figure 2**.

**Figure 2:**
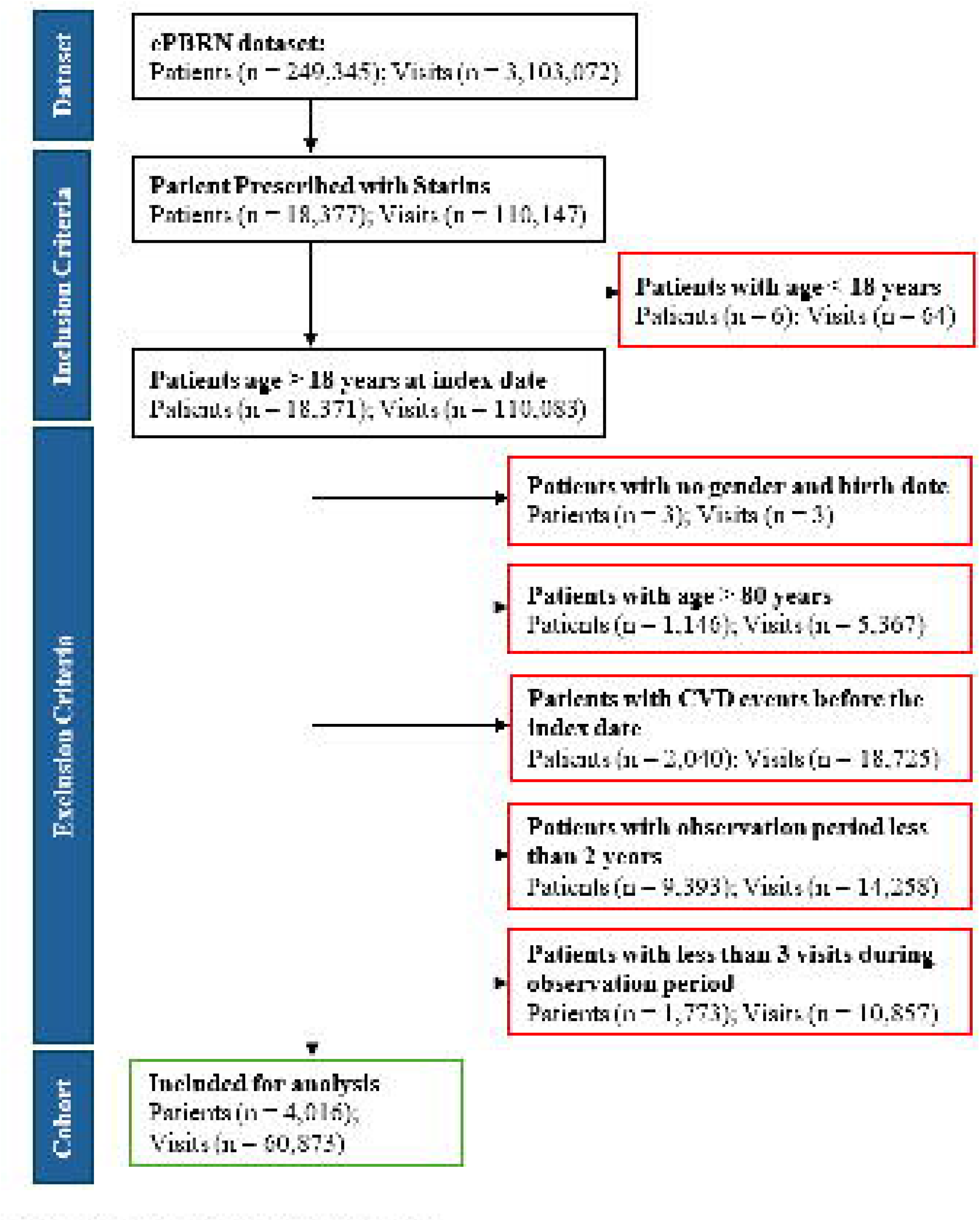
Selection of eligible patients. **Notes:** ePBRN: Electronic Practice Based Research Network; CVD: Cardiovascular disease

### 3.2 Descriptive characteristics

About 3.5% of patients were statin intolerant. The majority of the intolerant patients were aged 60–69 years (30.7%) and 50–59 years (28.6%), representing the two most prevalent age groups **(Table 1)**. More than half of the intolerant patients were female or other gender (57.1%). Most of the intolerant patients were non-Aboriginals and/or Torres Strait Islanders (90.0%) with the largest proportion from SEIFA IRSAD category 8 or 9 (61.4%). The employment status was not recorded for 60.0% of the statin intolerant patients. Nearly half of the intolerant patients were current smokers (46.4%), and the other half were non-smokers (45.0%). At the index date, 47.1% of intolerant patients were prescribed atorvastatin, and 68.6% received it at a moderate intensity dosage. Over the two years of observation period, there was no change in statin for 91.4% intolerant patients, and no change in statin intensity for 90.0% intolerant patients. About 66.4% patients were concurrently taking one additional medication alongside statin. Majority of the statin intolerant patients presented with one (42.1%) or no (40.0%) additional selected comorbidities.

**Table 1:** Characteristics of patients, stratified by statin tolerance status (N = 4,016)

| Variable | Total<br>(N = 4,016) | Statin intolerance status |  | P-value |
| --- | --- | --- | --- | --- |
|  |  | Intolerant<br>(n = 140) | Tolerant<br>(n = 3,876) |  |
| ***Socio-economic and demographic factors*** |  |  |  |  |
| Age in years [mean (SD)] | 58.70 (10.90) | 61.30 (10.5) | 58.60 (10.90) | < 0.01 |
| Age in years [n (%)] |  |  |  |  |
| < 30 | 22 (0.55) | < 5 | 22 (0.57) | 0.063 |
| 30 – 39 | 164 (4.08) | < 5 | 160 (4.13) |  |
| 40 – 49 | 608 (15.14) | 15 (10.71) | 593 (15.30) |  |
| 50 – 59 | 1,269 (31.60) | 40 (28.57) | 1,229 (31.71) |  |
| 60 – 69 | 1,232 (30.68) | 43 (30.71) | 1,189 (30.68) |  |
| ≥ 70 | 721 (17.95) | 38 (27.14) | 683 (17.62) |  |
| Gender |  |  |  |  |
| Male | 2,023 (50.37) | 60 (42.86) | 1,963 (50.64) | < 0.01 |
| Female or other | 1,993 (49.63) | 80 (57.14) | 1,913 (49.36) |  |
| Ethnicity |  |  |  |  |
| Non-ATSI | 3,038 (75.65) | 126 (90.00) | 2,912 (75.13) | < 0.01 |
| ATSI or not provided/<br>recorded<sup>1</sup> | 978 (24.35) | 14 (10.00) | 964 (24.87) |  |
| SEIFA IRSAD category (decile) |  |  |  |  |
| 1 | 726 (18.08) | 19 (13.57) | 707 (18.24) | < 0.001 |
| 2 | 407 (10.13) | < 5 | 403 (10.40) |  |
| 3 | 607 (15.11) | < 5 | 605 (15.61) |  |
| 4 | 1,073 (26.72) | 29 (20.71) | 1,044 (26.93) |  |
| 8 or 9 | 1,203 (29.96) | 86 (61.43) | 1,117 (28.82) |  |
| Employment status |  |  |  |  |
| Unemployed | 264 (6.57) | 8 (5.71) | 256 (6.60) | < 0.05 |
| Employed | 697 (17.36) | 17 (12.14) | 680 (17.54) |  |
| Retired and/or pensioner | 568 (14.14) | 31 (22.14) | 537 (13.85) |  |
| Not recorded <sup>1</sup> | 2,487 (61.93) | 84 (60.00) | 2,403 (62.00) |  |
| ***Health related factor*** |  |  |  |  |
| Smoking status |  |  |  |  |
| Non-smoker or ex-smoker | 1,901 (47.34) | 63 (45.00) | 1,838 (47.42) | 0.433 |
| Current smoker | 1,646 (40.99) | 65 (46.43) | 1,581 (40.79) |  |
| Not recorded <sup>1</sup> | 469 (11.68) | 12 (8.57) | 457 (11.79) |  |
| ***Statin related factors*** |  |  |  |  |
| Statin intensity at index date |  |  |  |  |
| High | 853 (21.24) | 37 (26.43) | 816 (21.05) | 0.311 |
| Moderate | 2,946 (73.36) | 96 (68.57) | 2,850 (73.53) |  |
| Low | 217 (5.40) | 7 (5.00) | 210 (5.42) |  |
| Statin type at index date |  |  |  |  |
| Atorvastatin | 1,887 (46.99) | 66 (47.14) | 1,821 (46.98) | 0.580 |
| Rosuvastatin | 1,255 (31.25) | 46 (32.86) | 1,209 (31.19) |  |
| Simvastatin | 681 (16.96) | 19 (13.57) | 662 (17.08) |  |
| Fluvastatin or Pravastatin | 193 (4.81) | 9 (6.43) | 184 (4.75) |  |
| Number of statin types |  |  |  |  |
| 1 | 3,585 (89.27) | 128 (91.43) | 3,457 (89.19) | 0.774 |
| ≥ 2 | 431 (10.73) | 12 (8.57) | 419 (10.81) |  |
| Change in statin intensity |  |  |  |  |
| No change | 3,514 (87.5) | 126 (90.00) | 3,388 (87.41) | 0.807 |
| High to Low or Moderate | 112 (2.79) | 4 (2.86) | 105 (2.79) |  |
| Moderate to High | 289 (7.20) | 9 (6.43) | 280 (7.22) |  |
| Moderate to Low | 27 (0.67) | < 5 | 26 (0.67) |  |
| Low to High | 9 (0.22) | < 5 | 9 (0.23) |  |
| Low to Moderate | 65 (1.62) | < 5 | 65 (1.68) |  |
| Number of prescribed medications<sup>2</sup> |  |  |  |  |
| 1<sup>3</sup> | 238 (5.93) | < 5 | 238 (6.14) | < 0.001 |
| 2 | 2,632 (65.54) | 93 (66.43) | 2,539 (65.51) |  |
| 3 | 1,008 (25.10) | 40 (28.57) | 968 (24.97) |  |
| 4 | 131 (3.26) | 5 (3.57) | 126 (3.25) |  |
| 5 | 7 (0.17) | < 5 | 5 (0.13) |  |
| Number of selected comorbidities<sup>4</sup> |  |  |  |  |
| None | 2,558 (63.70) | 56 (40.00) | 2,502 (64.55) | < 0.001 |
| 1 | 1,097 (27.32) | 59 (42.14) | 1,038 (26.78) |  |
| 2 | 333 (8.29) | 19 (13.57) | 314 (8.10) |  |
| ≥ 3 | 28 (0.70) | 6 (4.29) | 22 (0.57) |  |

**Notes:**
<sup>1</sup> Not recorded categories represent missing EHR information and should not be interpreted as patient-level clinical predictors;
<sup>2</sup> The categories for prescribed medications included anti-diabetic drugs, anti-hypertensive drugs, anti-coagulant drugs and one or more of other drugs;
<sup>3</sup> The number of prescribed medications = 1 are the people who are only on a statins;
<sup>4</sup> The categories for comorbidities included diabetes, hypertension, chronic respiratory disease, cancer, stroke, blood clots and Parkinson's disease;
ATSI: Aboriginals and Torres Strait Islanders; SEIFA IRSAD: Socio-Economic Indexes for Areas Index of Relative Socio-Economic Advantage and Disadvantage (with lower categories indicating greater disadvantage and higher categories indicating greater advantage)

### 3.3 Factors associated with statin intolerance

**Table 2** shows the univariate and multivariate logistic regression analysis for factors associated with statin intolerance. After adjusting for all other variables, higher odds of statin intolerance were observed among female or other gender patients (AOR 1.5, 95% CI 1.0 – 2.2), patients residing in SEIFA IRSAD category 8 or 9 (AOR 3.4, 95% CI 2.0 – 5.8), and patients with three or more of the selected comorbidities (AOR 6.4, 95% CI 2.2 – 18.3), compared with their respective reference categories.

**Table 2:** Univariate and multivariate logistic regression analysis for factors associated with statin intolerance (*N* = 4,016)

| Factors | n (%) | Unadjusted Odds ratio<br>(95% CI) | p-value | Adjusted Odds ratio<br>(95% CI) | p-value |
| --- | --- | --- | --- | --- | --- |
| ***Socio-economic and demographic factors*** |  |  |  |  |  |
| Age (years) |  |  |  |  |  |
| ≤ 65 | 2,883 (71.79) | Reference |  | Reference |  |
| > 65 | 1,133 (28.21) | 1.73 (1.23 – 2.45) ** | < 0.01 | 1.19 (0.76 – 1.84) | 0.446 |
| Gender |  |  |  |  |  |
| Male | 2,023 (50.37) | Reference |  | Reference |  |
| Female or other | 1,993 (49.63) | 1.37 (0.97 – 1.92) | 0.071 | 1.47 (0.99 – 2.18) | 0.055 |
| SEIFA IRSAD category (decile) |  |  |  |  |  |
| 1 | 726 (18.08) | Reference |  | Reference |  |
| 2 | 407 (10.13) | 0.37 (0.12 – 1.09) | 0.072 | 0.49 (0.16 – 1.49) | 0.210 |
| 3 | 607 (15.11) | 0.12 (0.03 – 0.53) ** | < 0.01 | 0.19 (0.04 – 0.86) * | < 0.05 |
| 4 | 1,073 (26.72) | 1.03 (0.587 – 1.86) | 0.912 | 1.17 (0.63 – 2.16) | 0.616 |
| 8 or 9 | 1,203 (29.96) | 2.86 (1.73 – 4.75) *** | < 0.001 | 3.41 (2.00 – 5.82) *** | < 0.001 |
| Employment status |  |  |  |  |  |
| Unemployed | 264 (6.57) | Reference |  | Reference |  |
| Employed | 697 (17.36) | 0.53 (0.28 – 1.02) | 0.058 | 0.82 (0.38 – 1.79) | 0.625 |
| Retired and/or pensioner | 568 (14.14) | 1.15 (0.64 – 2.05) | 0.637 | 1.50 (0.75 – 2.99) | 0.249 |
| Not recorded <sup>1</sup> | 2,487 (61.93) | N/A |  | N/A |  |
| ***Health related factor*** |  |  |  |  |  |
| Smoking status |  |  |  |  |  |
| Non-smoker or ex-smoker | 1,891 (47.09) | Reference | Reference |  |  |
| Current smoker | 1,646 (40.99) | 1.17 (0.82 – 1.65) | 0.388 | 1.27 (0.87 – 1.86) | 0.210 |
| Not recorded <sup>1</sup> | 469 (11.68) | N/A |  | N/A |  |

| ***Statin related factors*** |  |  |  |  |  |
| --- | --- | --- | --- | --- | --- |
| Statin intensity at index date |  |  |  |  |  |
| Moderate | 2,946 (73.36) | *Reference* |  | *Reference* |  |
| High | 853 (21.24) | 1.35 (0.91 – 1.98) | 0.132 | 1.37 (0.86 – 2.16) | 0.183 |
| Low | 217 (5.40) | 0.99 (0.45 – 2.16) | 0.979 | 1.17 (0.44 – 3.08) | 0.753 |
| Statin type at index date |  |  |  |  |  |
| Atorvastatin | 1,887 (46.99) | *Reference* |  | *Reference* |  |
| Rosuvastatin | 1,255 (31.25) | 1.05 (0.72 – 1.54) | 0.804 | 1.00 (0.66 – 1.53) | 0.985 |
| Simvastatin | 681 (16.96) | 0.79 (0.47 – 1.33) | 0.377 | 0.77 (0.42 – 1.40) | 0.390 |
| Fluvastatin or Pravastatin | 193 (4.81) | 1.35 (0.66 – 2.75) | 0.410 | 1.50 (0.62 – 3.66) | 0.368 |
| Number of statin types |  |  |  |  |  |
| 1 | 3,585 (89.27) | *Reference* |  | *Reference* |  |
| ≥ 2 | 431 (10.73) | 0.77 (0.42 – 1.41) | 0.402 | 1.02 (0.52 – 2.01) | 0.947 |
| Change in statin intensity |  |  |  |  |  |
| No change | 3,514 (87.5) | *Reference* |  | *Reference* |  |
| Low to moderate or high | 74 (1.84) | 0.00 (N/A) | 0.975 | 0.00 (N/A) | 0.983 |
| Moderate to low | 27 (0.67) | 1.03 (0.14 – 7.69) | 0.974 | 1.39 (0.17 – 11.43) | 0.759 |
| Moderate to high | 289 (7.20) | 0.86 (0.43 – 1.72) | 0.677 | 1.13 (0.54 – 2.35) | 0.749 |
| High to low or moderate | 112 (2.79) | 1.00 (0.36 – 2.75) | 0.994 | 0.96 (0.31 – 2.96) | 0.939 |
| Number of prescribed medications <sup>2</sup> |  |  |  |  |  |
| 1 <sup>3</sup> | 238 (5.93) | *Reference* |  | *Reference* |  |
| 2 | 2,632 (65.54) | 4236052.69 (N/A) | 0.971 | 2984129.86 (N/A) | 0.970 |
| 3 | 1,008 (25.10) | 4778875.76 (N/A) | 0.971 | 2976719.41 (N/A) | 0.970 |
| 4 | 131 (3.26) | 4589237.83 (N/A) | 0.971 | 2491592.33 (N/A) | 0.971 |
| 5 | 7 (0.17) | 46259517.35 (N/A) | 0.967 | 21564768.24 (N/A) | 0.966 |

| Number of selected comorbidities <sup>4</sup> |  |  |  |  |  |
| --- | --- | --- | --- | --- | --- |
| None | 2,558 (63.70) | *Reference* |  | *Reference* |  |
| 1 | 1,097 (27.32) | 2.54 (1.75 – 3.69) *** | < 0.001 | 1.48 (1.00 – 2.20) | 0.051 |
| 2 | 333 (8.29) | 2.70 (1.59 – 4.61) *** | < 0.001 | 1.37 (0.77 – 2.45) | 0.286 |
| ≥ 3 | 28 (0.70) | 12.19 (4.75 – 31.23) *** | < 0.001 | 6.37 (2.22 – 18.26) ** | < 0.01 |

**Notes:**
<sup>1</sup> Not recorded categories represent missing EHR information and should not be interpreted as patient-level clinical predictors;
<sup>2</sup> The categories for prescribed medications included anti-diabetic drugs, anti-hypertensive drugs, anti-coagulant drugs and one or more of other drugs;
<sup>3</sup> The number of prescribed medications = 1 are the people who are only on statins;
<sup>4</sup> The categories for comorbidities included diabetes, hypertension, chronic respiratory disease, cancer, stroke, blood clots and Parkinson's disease;
ATSI: Aboriginals and Torres Strait Islanders; SEIFA IRSAD: Socio-Economic Indexes for Areas Index of Relative Socio-Economic Advantage and Disadvantage (with lower categories indicating greater disadvantage and higher categories indicating greater advantage); CI: Confidence interval; N/A: Not Applicable

### 3.4 Predictors of statin intolerance in the final model

The logistic regression of the overall model met all key assumptions. The overall regression model showed adequate fit (Hosmer-Lemeshow test: p = 0.75) and acceptable discriminative ability (AUC = 0.78). The details on the assumptions of the regression model are shown in **Supplement 3**.

Backward stepwise model selection using AIC was performed to identify a parsimonious final model containing the key predictors of statin intolerance **(Table 3)**. The final model had a lower AIC than the overall model, and the likelihood ratio test indicated that removal of the excluded variables did not significantly worsen model fit. The final model showed acceptable discriminative ability. Although its AUC was significantly lower than that of the overall model, the absolute difference was small. The estimates of the final model are shown in **Supplements 4 and 5**. **Supplement 6** shows the components of the overall regression model and the final model and their explanations.

**Table 3:**
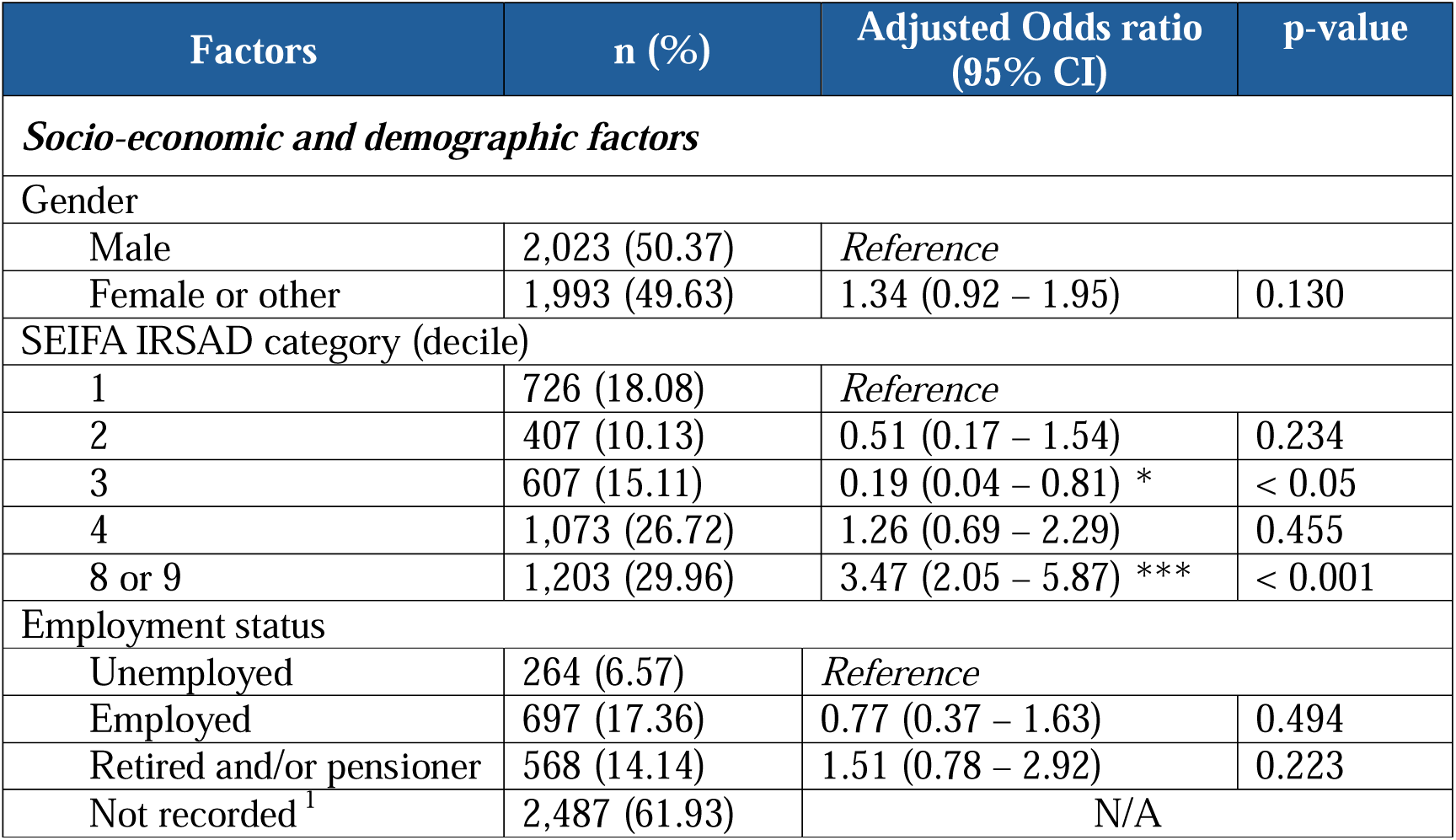
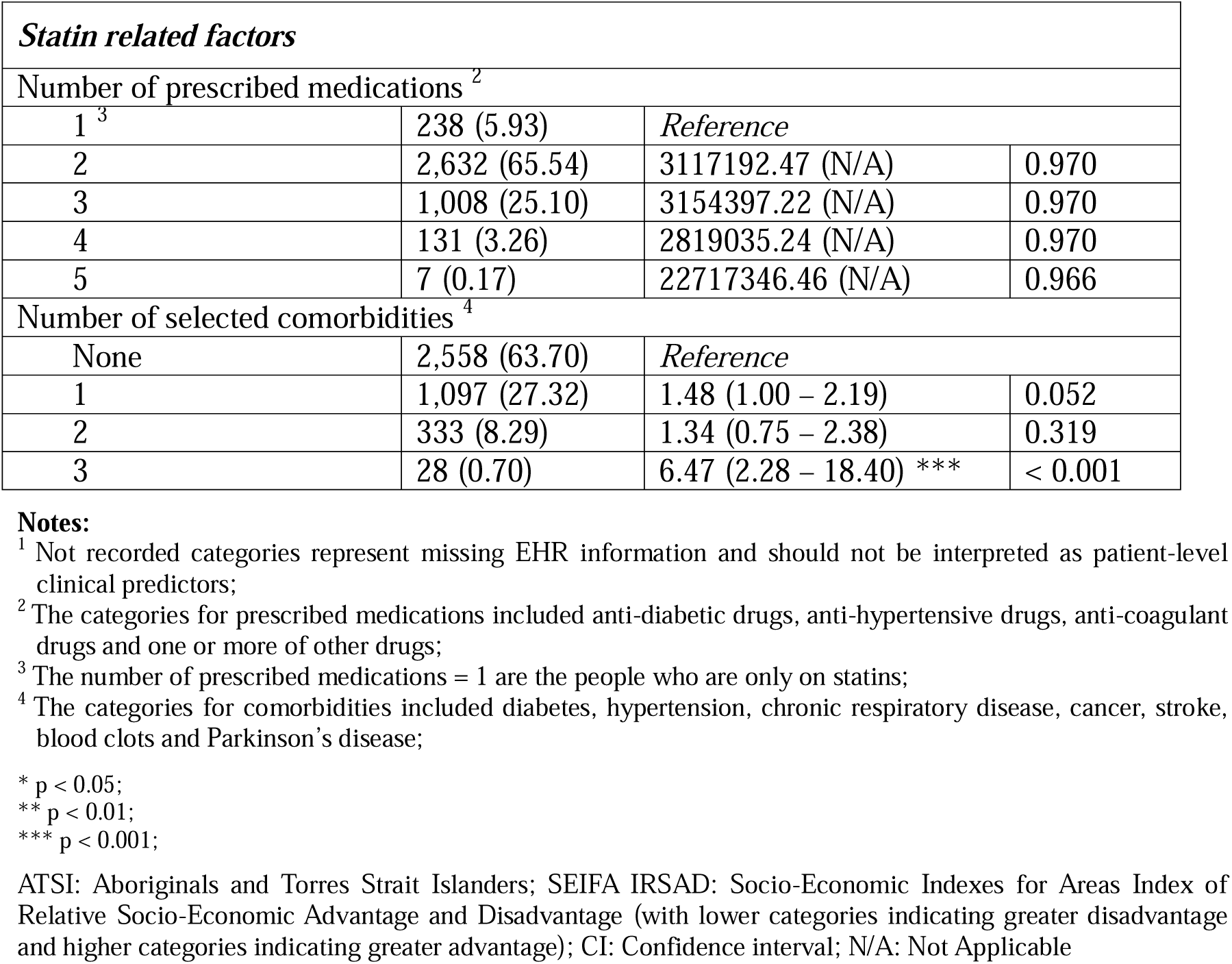
Predictors of statin intolerance identified in the final multivariate logistic regression model.

| Factors | n (%) | Adjusted Odds ratio<br>(95% CI) | p-value |
| --- | --- | --- | --- |
| ***Socio-economic and demographic factors*** |  |  |  |
| Gender |  |  |  |
| Male | 2,023 (50.37) | *Reference* |  |
| Female or other | 1,993 (49.63) | 1.34 (0.92 – 1.95) | 0.130 |
| SEIFA IRSAD category (decile) |  |  |  |
| 1 | 726 (18.08) | *Reference* |  |
| 2 | 407 (10.13) | 0.51 (0.17 – 1.54) | 0.234 |
| 3 | 607 (15.11) | 0.19 (0.04 – 0.81) * | < 0.05 |
| 4 | 1,073 (26.72) | 1.26 (0.69 – 2.29) | 0.455 |
| 8 or 9 | 1,203 (29.96) | 3.47 (2.05 – 5.87) *** | < 0.001 |
| Employment status |  |  |  |
| Unemployed | 264 (6.57) | *Reference* |  |
| Employed | 697 (17.36) | 0.77 (0.37 – 1.63) | 0.494 |
| Retired and/or pensioner | 568 (14.14) | 1.51 (0.78 – 2.92) | 0.223 |
| Not recorded<sup>1</sup> | 2,487 (61.93) | N/A |  |

| ***Statin related factors*** |  |  |  |
| --- | --- | --- | --- |
| Number of prescribed medications <sup>2</sup> |  |  |  |
| 1 <sup>3</sup> | 238 (5.93) | *Reference* |  |
| 2 | 2,632 (65.54) | 3117192.47 (N/A) | 0.970 |
| 3 | 1,008 (25.10) | 3154397.22 (N/A) | 0.970 |
| 4 | 131 (3.26) | 2819035.24 (N/A) | 0.970 |
| 5 | 7 (0.17) | 22717346.46 (N/A) | 0.966 |
| Number of selected comorbidities <sup>4</sup> |  |  |  |
| None | 2,558 (63.70) | *Reference* |  |
| 1 | 1,097 (27.32) | 1.48 (1.00 – 2.19) | 0.052 |
| 2 | 333 (8.29) | 1.34 (0.75 – 2.38) | 0.319 |
| 3 | 28 (0.70) | 6.47 (2.28 – 18.40) *** | < 0.001 |

**Notes:**
<sup>1</sup> Not recorded categories represent missing EHR information and should not be interpreted as patient-level clinical predictors;
<sup>2</sup> The categories for prescribed medications included anti-diabetic drugs, anti-hypertensive drugs, anti-coagulant drugs and one or more of other drugs;
<sup>3</sup> The number of prescribed medications = 1 are the people who are only on statins;
<sup>4</sup> The categories for comorbidities included diabetes, hypertension, chronic respiratory disease, cancer, stroke, blood clots and Parkinson's disease;
\* p < 0.05;
\*\* p < 0.01;
\*\*\* p < 0.001;
ATSI: Aboriginals and Torres Strait Islanders; SEIFA IRSAD: Socio-Economic Indexes for Areas Index of Relative Socio-Economic Advantage and Disadvantage (with lower categories indicating greater disadvantage and higher categories indicating greater advantage); CI: Confidence interval; N/A: Not Applicable

## 4. Discussion

In this study, 3.49% of patients in the ePBRN dataset were identified as statin intolerant, which is lower than the reported prevalence of statin intolerance in Australia (5–15% of patients) (4, 5). Global and Australian estimates of statin intolerance vary considerably according to the definitions and methods used to identify statin intolerance (6). Statin intolerance generally includes adverse effects attributed to statin therapy, particularly statin associated muscle symptoms, and the inability to maintain statin therapy at guideline directed doses despite attempts at rechallenge (41). Estimates may also vary according to the distinction between complete and partial statin intolerance and the CK threshold applied (4, 5, 42). The relatively low prevalence observed in this study may partly reflect the restrictive operational definition used, under which patients with muscle symptoms without elevated CK were not classified as statin intolerant. As statin associated muscle symptoms can occur without CK elevation, some clinically relevant cases may not have been identified. Therefore, the prevalence observed in this study should be interpreted in relation to the operational EHR based definition used rather than as an estimate of all clinical presentations of statin intolerance. The low prevalence may also be partly explained by the patient selection criteria (11). Only RACGP active patients were included, which may have excluded some patients who experienced symptoms and discontinued statin therapy without returning to their GP, or who returned after the two years observation period. In addition, patients may have received care from general practices or other healthcare providers outside the ePBRN network during the observation period. Statin associated symptoms, CK measurements, treatment changes, or discontinuation occurring outside participating practices may therefore not have been captured in the available EHR data. This incomplete capture of care may have resulted in some cases of statin intolerance not being identified and may have contributed to the observed lower observed prevalence.

This study found that gender, area level socioeconomic advantage and disadvantage, and comorbidities were associated with statin intolerance. Several studies have reported that females experience higher rates of statin intolerance or SAMS than males, possibly because of differences in muscle physiology, hormonal factors, and/or comorbidity profiles (7, 9, 19, 43). This aligns with the findings of this study, which identified an association between female gender and higher odds of statin intolerance than in males. However, such findings are variable and may depend on the population, study design, and definitions used to identify statin intolerance. The heterogeneity in the findings highlights the need for a standardized approach to identifying statin intolerance using EHR data through phenotyping algorithms. The literature suggests that statin intolerance is multifactorial, and several other sociodemographic factors may influence its reporting, identification, estimation, and management (9, 19). Although socioeconomic status (SES) is not consistently identified as a predictor of statin intolerance, some studies have suggested that patients from more socioeconomically disadvantaged areas may experience higher rates of intolerance than those from less disadvantaged areas (41). However, in contrast to these findings, this study found that patients from relatively advantaged areas (SEIFA IRSAD category 8 or 9) had about 240% higher odds of intolerance, compared with patients from more disadvantaged areas (SEIFA IRSAD category 1). This association may reflect differences in healthcare utilization, symptom reporting, or documentation rather than a direct effect of socioeconomic circumstances on statin intolerance. Studies have also suggested that a greater burden of comorbidities may increase the likelihood of experiencing symptoms associated with statin intolerance (44). Employment status and smoking status were included in the regression analysis following multiple imputation of missing information. However, the substantial proportion of missing employment data (61.93%) warrants caution when interpreting any association involving this variable, as the imputed values depend on assumptions about the missing data mechanism and the information available for imputation.

The findings of this study may have clinical implications for identifying patients who may require closer assessment of statin intolerance. Characteristics associated with statin intolerance, including female gender, area level socioeconomic circumstances, and a higher burden of comorbidities, may help clinicians identify patients who could benefit from further assessment after statin initiation. These factors are not diagnostic on their own but may support earlier review of symptoms and treatment, including dose modification, use of an alternative statin, or rechallenge where appropriate. The findings may also be relevant to Australian primary care, although their generalizability should be considered carefully. The ePBRN data were obtained from general practices in South Western Sydney, which has a culturally and socioeconomically diverse population. Differences in patient characteristics, healthcare access, prescribing practices, and EHR documentation across Australia may influence the observed associations, particularly those involving socioeconomic circumstances. Further validation in other Australian populations and settings is therefore needed before the findings can be considered broadly generalizable.

### Limitations

This study used prescription data from the ePBRN Linked Dataset from general practices. Statin intolerance lacks a standardized taxonomy and has varying definitions, which may influence case identification. The operational definition used in this study was restrictive, as patients with muscle symptoms without elevated CK were not classified as statin intolerant. As statin associated muscle symptoms can occur with normal CK levels and may contribute to statin reduction or discontinuation, some clinically relevant cases may have been misclassified. A sensitivity analysis using a broader definition of statin intolerance was not performed. Consequently, it was not possible to determine whether inclusion of patients with muscle symptoms without elevated CK would have changed the identified predictors or the predictive performance of the model. The findings should therefore be interpreted in relation to the operational EHR based definition used in this study rather than all clinical presentations of statin associated muscle symptoms.

The dataset also contained substantial missing information for some sociodemographic and clinical variables, particularly employment status. To address missing information in employment status (61.93%) and smoking status (11.68%), MICE was performed rather than retaining “not recorded” as a separate category in the regression analysis. Although this approach allowed the available study sample to be retained, it relied on the assumption that the missing data were missing at random, conditional on the variables included in the imputation model. This assumption could not be verified from the available data. Given the high proportion of missing employment information, the imputed values may not fully represent the underlying employment characteristics of the study population, particularly if missingness was related to factors not captured in the EHR data. Consequently, associations involving employment status should be interpreted cautiously, as they may be influenced by uncertainty in the imputation process and unmeasured factors associated with missing documentation. The ePBRN data were limited to information recorded within participating general practices, and care received outside the ePBRN network may not have been captured. Although all available EHR records before the index date were searched for evidence of prior statin use, patients may have received statin prescriptions from healthcare providers outside the ePBRN network before their first recorded statin prescription in the dataset. Therefore, the index date represents the first recorded statin prescription within the available ePBRN data and may not correspond to the patient’s first ever use of a statin. Consequently, relevant symptoms, investigations, or changes to statin therapy occurring elsewhere may have been missed, potentially resulting in misclassification of statin intolerance.

The assessment of comorbidities was restricted to the predefined categories included in the analysis and did not capture all possible comorbid conditions. Patients may have had other conditions that were not represented in the comorbidity count, potentially underestimating their overall comorbidity burden. This may have influenced the observed association between comorbidity burden and statin intolerance and resulted in residual confounding from conditions not included in the analysis.

Although backward AIC model selection was performed across the multiple imputed datasets, the selection of the most frequently occurring reduced model does not fully account for uncertainty in variable selection across imputations. In addition, the predictive performance of the regression models was assessed using the same dataset employed for model development, and external validation was not performed. Therefore, the reported discriminatory performance may be optimistic and may not reflect model performance in other populations. The use of a single data source (the ePBRN dataset) and incomplete recording of sociodemographic variables also limits the generalizability of the findings. Generalizability may be further limited by excluding patients who experienced CVD events before the index date or during the two years observation period, which may have introduced selection bias by excluding patients at higher risk of CVD events.

## 5. Conclusion

This study used the ePBRN dataset from South-western Sydney and identified statin intolerance in 3.5% of patients. Female or other gender, residence in SEIFA IRSAD category 8 or 9, and having three or more of the selected comorbidities were associated with about 50%, 240%, and 540% higher odds of statin intolerance, respectively, in the adjusted analysis. However, these findings should be interpreted in the context of the study population, the operational definition of statin intolerance, and the limitations of routinely collected EHR data. Although multiple imputation was used to address missing employment and smoking information, the substantial proportion of missing employment data warrants caution when interpreting the findings. Further validation in other populations and settings is required to assess the generalizability of the identified predictors.

## Data Availability

The corresponding author will make the data available upon request.

## Author contributions

1. SR and JJ contributed to the conceptualization of this study.
2. JJ contributed to the funding acquisition
3. JR, KAR, STL and JJ contributed to the resources and supervision of this study
4. JR, KAR, STL and JJ contributed to the validation
5. All authors contributed to the reviewing and editing of this study
6. SR and JJ contributed to data curation
7. SR contributed to the formal analysis and the original drafting of this study
8. SR contributed to the methodology of the study

## Data sharing statement

The corresponding author will make the data available upon request.

## Acknowledgements

We would like to thank the ePBRN Primary Care Health Informatics Working Group of the Secure Research Environment for Digital Health (SREDH) Consortium (https://www.sredhconsortium.org, accessed on 7 October 2024) for their assistance with access to the ePBRN dataset to investigate the findings from this review.

