## Supplement for "Predictors of statin intolerance among statin users in primary care using real-world data"

### **Supplement 1: List of statins included in the ePBRN dataset**

| **Best practice** | **Medical Director** |
| --- | --- |
| 1. Atorvastatin 2. Amlodipine besylate 3. Amlodipine 4. Atorvachol 5. Atozet 6. Cadatin 7. Cadivast 8. Caduet 9. Cavstat 10. Cholstat 11. Crestor 12. Crosuva 13. Ezalo 14. Fluvastatin 15. Lescol 16. Lipex 17. Lipitor 18. Lipostat 19. Liprachol 20. Lorstat 21. Pravastatin 22. Pravastatin sodium 23. Pravachol% 24. Pravastat 25. Rosuvastatin 26. Rosuzet 27. Simvastatin 28. Simvacor 29. Simvahexal 30. Simvar 31. Torvastat 32. Trovas 33. Vastin 34. Visacor 35. Vytorin 36. Zimstat 37. Zocor 38. Zeklen | 1. Atorvastatin 2. Pravastatin 3. Rosuvastatin 4. Simvastatin 5. Atorvachol 6. Atozet 7. Cadatin 8. Cadivast 9. Caduet 10. Cavstat 11. Crestor 12. Crosuva 13. Ezalo 14. Fluvastatin 15. Lescol 16. Lescol 17. Lipex 18. Lipitor 19. Lorstat 20. Pravachol 21. Simvacor 22. Simvahexal 23. Simvar 24. Torvastat 25. Trovas 26. Vastin 27. Zimstat 28. Zocor |

**Supplement 2: Statin induced dose-based reductions of LDL-C in Australia**

| High-intensity statins (> 50% reduction in LDL-C) | Moderate-intensity statins (30–49% reduction in LDL-C) | Low-intensity statins (< 30% reduction in LDL-C) |
| --- | --- | --- |
| - Atorvastatin 40–80 mg - Rosuvastatin 20–40 mg | - Atorvastatin 10–20 mg - Fluvastatin 80 mg - Pravastatin 40–80 mg - Rosuvastatin 5–10 mg - Simvastatin 20–80 mg | - Fluvastatin 20–40 mg - Pravastatin 10–20 mg - Simvastatin 5–10 mg |

LDL-C: Low-density lipoprotein cholesterol

**Supplement 3: Assumptions of overall regression model**

**Binary outcome:** The outcome of interest was binary (ie. tolerant or intolerant)

**Independent observation:** Satisfies the independence of observation.

**Multicollinearity:**

| **Factors** | **Overall regression model** | | |
| --- | --- | --- | --- |
|  | **GVIF** | **Df** | **GVIF^(1/(2*Df))** |
| Age | 1.36 | 1 | 1.17 |
| Gender | 1.20 | 1 | 1.09 |
| SEIFA IRSAD category (decile) | 1.20 | 4 | 1.02 |
| Employment status | 1.71 | 2 | 1.14 |
| Smoking status | 1.11 | 1 | 1.05 |
| Statin intensity at index date | 1.96 | 2 | 1.18 |
| Statin type at index date | 1.84 | 3 | 1.11 |
| Number of statin types | 1.22 | 1 | 1.10 |
| Change in statin intensity | 1.37 | 4 | 1.04 |
| Number of prescribed medications | 1.24 | 4 | 1.03 |
| Number of selected comorbidities | 1.26 | 3 | 1.04 |

GVIF: Generalized Variance Inflation Factor; Df: Degree of freedom

Multicollinearity among the independent variables included in the overall multivariate logistic regression model was assessed using the generalized variance inflation factor (GVIF). The GVIF values ranged from 1.11 to 1.96, while the adjusted GVIF values, calculated as GVIF^(1/(2*Df)), ranged from 1.02 to 1.18. These values were close to 1, indicating no evidence of substantial multicollinearity among the predictors included in the model. Although some association between the number of prescribed medications and comorbidities was expected, particularly for diabetes and hypertension, the two variables captured different aspects of patient characteristics. The variable of prescribed medication included prescriptions for antidiabetic, antihypertensive, anticoagulant and other medications, whereas the variable of comorbidities represented seven selected conditions. Medication use may also reflect preventive treatment or conditions not captured by the selected comorbidity categories, while a recorded comorbidity may not necessarily correspond to a prescription in the available EHR data. The adjusted GVIF values for the number of prescribed medications (1.03) and selected comorbidities (1.04) were low, suggesting that these variables did not exhibit substantial multicollinearity with the other predictors in the model, despite their potential clinical association.

**Goodness of fit:** The Hosmer–Lemeshow goodness of fit test did not indicate a statistically significant difference between the observed and the predicted outcomes (p-value = 0.7541), suggesting no evidence of poor model fit.

**Area under the curve:** The overall regression model demonstrated acceptable discriminatory ability, with an area under the receiver operating characteristic curve (AUC) of 0.783, indicating that the model could reasonably distinguish between patients with and without statin intolerance.

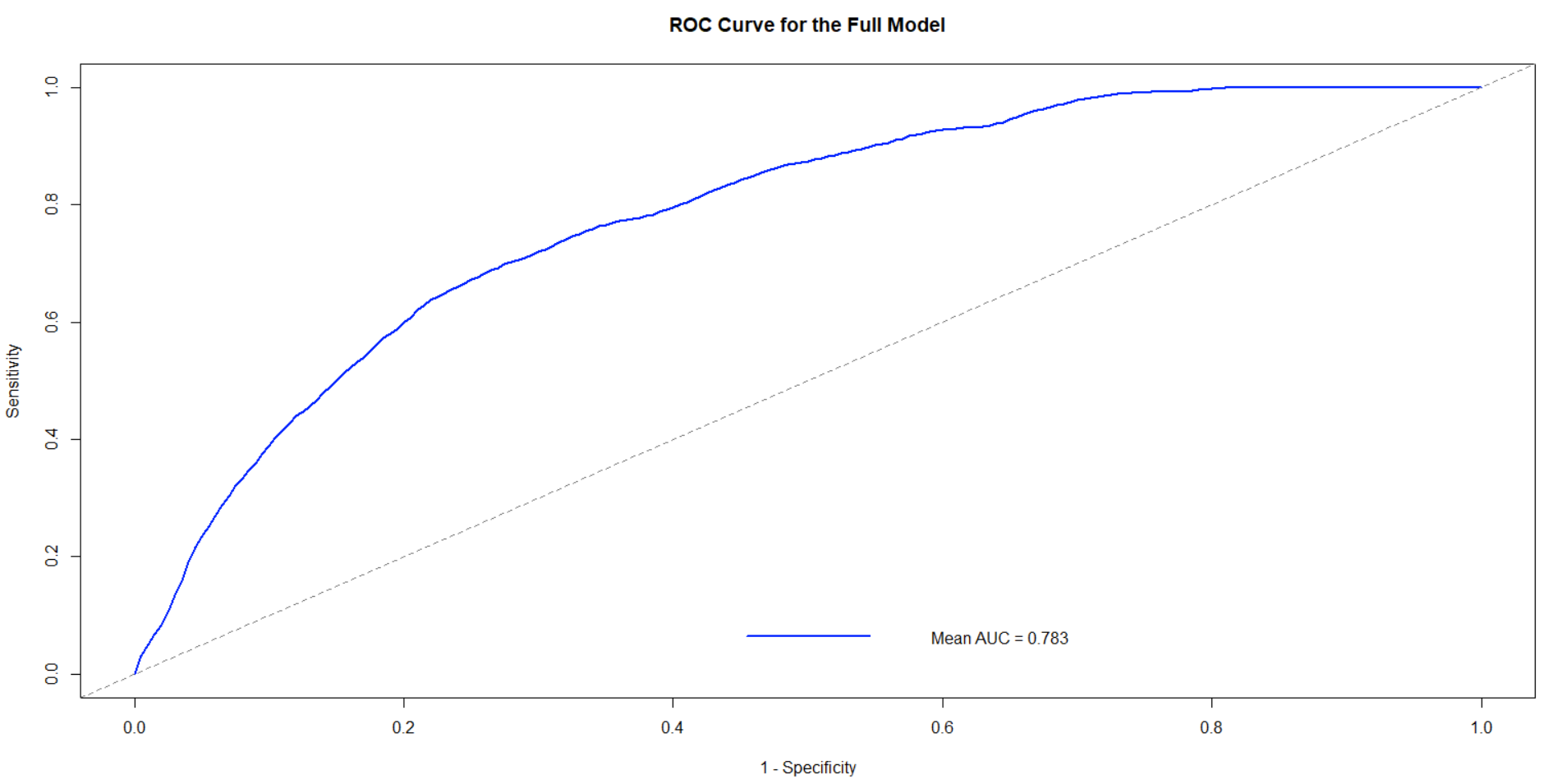

**Supplement 4: AIC of overall regression model vs final model**

| **Model** | **Df** | **Mean AIC** | **P-value** | **Interpretation** |
| --- | --- | --- | --- | --- |
| Overall regression model | 26 | 1120.933 | 0.8409 | Lower AIC of final model indicate a better balance between model fit and complexity. The likelihood ratio test showed no statistically significant difference between the two models, indicating that removal of the excluded variables did not significantly worsen the model fit. Therefore, the more parsimonious final model was preferred. |
| Final model | 14 | 1109.819 |  |  |

AIC: Akaike Information Criterion; Df: Degree of freedom

**Supplement 5: AUC of overall regression model vs final model**

| **Model** | **AUC** | **P-value** | **Interpretation** |
| --- | --- | --- | --- |
| Overall regression model | 0.7827 | 0.00717 | The overall regression model demonstrated slightly better discrimination than the final model. Although the difference in AUC was statistically significant, the absolute difference was small. |
| Final model | 0.7677 |  |  |

AUC: Area under the curve; Df: Degree of freedom

**Supplement 6: Model predictors of intolerance**

| **Model** | **Variable** | **Description** |
| --- | --- | --- |
| **Model 1 (overall regression model) and model 2 (final model)** | Gender | Gender as reported at index date |
|  | SEIFA IRSAD category (decile) | Decile of the Index of Relative Socio-economic Advantage and Disadvantage (IRSAD) for within Australia published in 2021 ^1^ |
|  | Employment status | Self-reported |
|  | Number of prescribed medications ^2^ | Number of medicine/drug classes prescribed to patient, as found in the dataset |
|  | Number of selected comorbidities ^3^ | Number of other comorbid conditions of the patient, as found in the dataset |
| **Model 1 (overall regression model) only** | Age | Age at index date (date of first statin prescription) |
|  | Smoking status | Self-reported |
|  | Statin intensity at index date | The intensity of statin prescribed at index date |
|  | Statin type at index date | The statin type prescribed at index date |
|  | Number of statin types | The number of statin types prescribed during the period between index date and last visit date in two years after the index date |
|  | Change in statin intensity | Change in the prescribed statin intensity from index date to last visit date in two years after the index date |

^1^ SEIFA IRSAD category (decile) was based on the Australian Bureau of Statistics (ABS) Socio-Economic Indexes for Areas (SEIFA), Australia, retrieved from: <https://www.abs.gov.au/statistics/people/people-and-communities/socio-economic-indexes-areas-seifa-australia/latest-release>;

^2^ The categories for prescribed medications included anti-diabetic drugs, anti-hypertensive drugs, anti-coagulant drugs and others drugs;

^3^ The categories for comorbidities included diabetes, hypertension, chronic respiratory disease, cancer, stroke, blood clots and Parkinson’s disease
